## Supplementary Figures for "Case-control investigation of invasive Salmonella disease in Malawi reveals no evidence of environmental or animal transmission of invasive strains, and supports human to human transmission"

### Slide 1
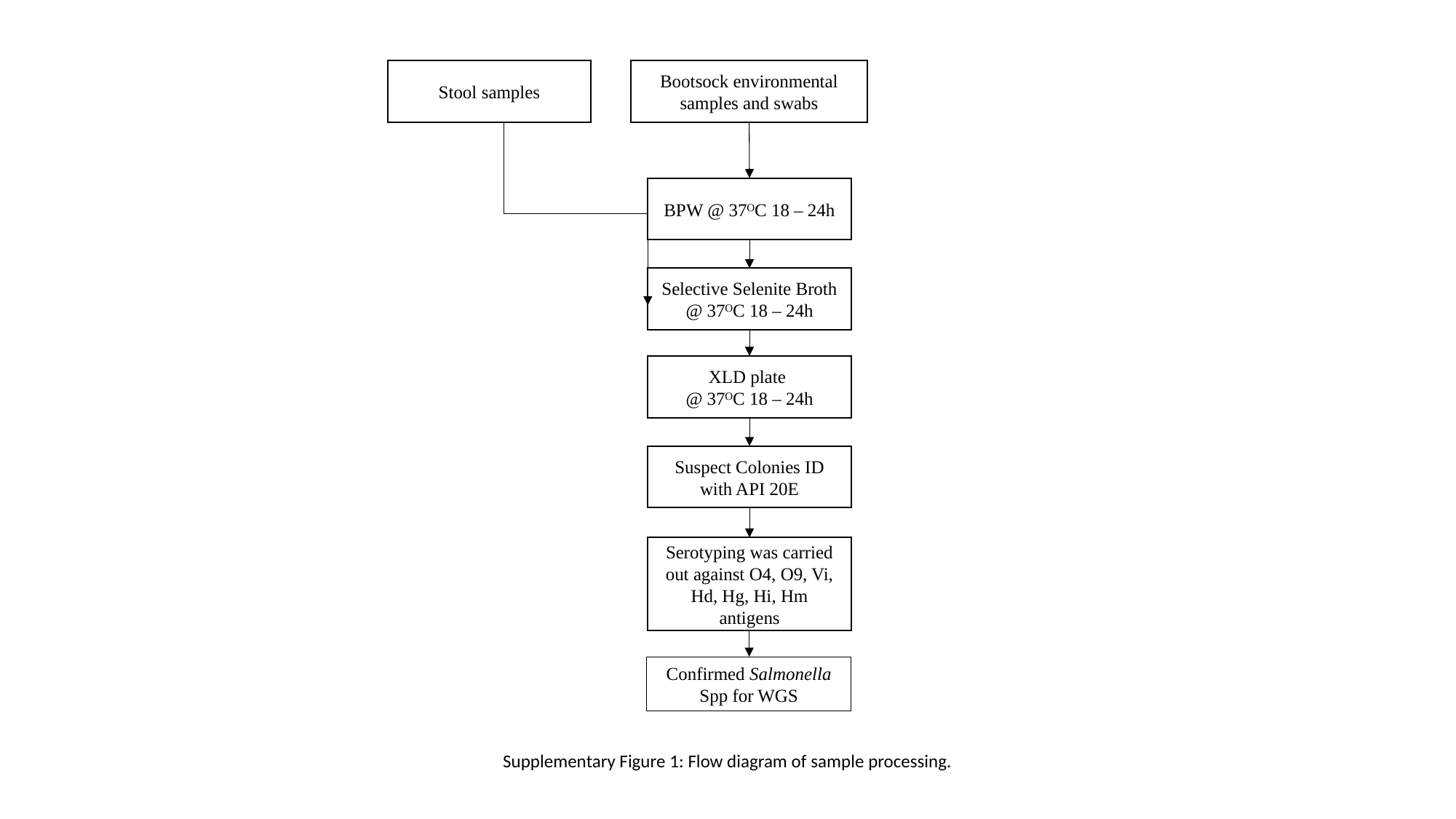

Stool samples
Bootsock environmental samples and swabs
BPW @ 37ᴼC 18 – 24h
Selective Selenite Broth @ 37ᴼC 18 – 24h
XLD plate
@ 37ᴼC 18 – 24h
Suspect Colonies ID with API 20E
Serotyping was carried out against O4, O9, Vi, Hd, Hg, Hi, Hm antigens
Confirmed Salmonella Spp for WGS
Supplementary Figure 1: Flow diagram of sample processing.
